# Evaluating diagnostic accuracies of Panbio^™^ COVID-19 rapid antigen test and RT-PCR for the detection of SARS-CoV-2 in Addis Ababa, Ethiopia using Bayesian Latent-Class Models (BLCM)

**DOI:** 10.1101/2022.04.25.22274285

**Authors:** Abay Sisay, Sonja Hartnack, Abebaw Tiruneh, Yasin Desalegn, Abraham Tesfaye, Adey Feleke Desta

**Affiliations:** Department of Medical Laboratory Sciences, College of Health Sciences, Addis Ababa University, Addis Ababa, Ethiopia; Department of Microbial, Cellular and Molecular Biology, College of Natural and Computational Sciences, Addis Ababa University, Addis Ababa, Ethiopia; Section of Epidemiology, Vetsuisse Faculty, University of Zurich, Zurich, Switzerland; Addis Ababa Public Health Research and Emergency Management Laboratory, Addis Ababa Health Bureau, Addis Ababa, Ethiopia; Diagnostic Unit, Center for Innovative Drug Development and Therapeutic Trials for Africa, CDT-Africa, Addis Ababa, Ethiopia

**Author notes:** These authors contributed equally to this work (Abay Sisay, Sonja Hartnack).

**Keywords:** diagnostic sensitivity, diagnostic specificity, no gold standard, rapid diagnostic tests

## Abstract

**Background:** Rapid diagnostics are vital for curving the transmission and control of COVID-19 pandemic. Although many commercially available antigen-based rapid diagnostic tests (Ag-RDTs) for the detection of SARS-CoV-2 are recommended by the WHO, their diagnostic performance has not yet been assessed in Ethiopia. So far, the vast majority of studies assessing diagnostic accuracies of rapid antigen tests considered RT-PCR as a gold standard, which inevitably leads to bias when RT-PCR is not 100% sensitive and specific. Thus, this study aimed to evaluate the diagnostic performance of Panbio ™ COVID-19 rapid antigen test jointly together with the RT-PCR for the detection of SARS-CoV-2.

**Methods:** A prospective cross-sectional study was done from July to September 2021 in Addis Ababa, Ethiopia, during the third wave of the pandemic involving two health centers and two hospitals. Diagnostic sensitivity and specificity of Panbio™ rapid antigen test and RT-PCR were obtained using Bayesian Latent-Class Models (BLCM).

**Results:** 438 COVID-19 presumptive clients were enrolled, 239 (54.6%) were females, of whom 196 (44.7%) had a positive RT-PCR and 158 (36.1%) were Ag-RDT positive. The Ag-RDT and RT-PCR had a sensitivity (95% CrI) of 99.6 (98.4-100), 89.3 (83.2-97.6) and specificity (95% CrI) of 93.4 (82.3 - 100), 99.1 (97.5-100) respectively. Most of the study participants, 318 (72.6) exhibited COVID-19 symptoms and the most reported was cough 191 (43.6).

**Conclusion:** The diagnostic performance of Panbio™ COVID-19 Ag RDT is coherent with the WHO established criteria of having a sensitivity ≥80% for Ag-RDTs. Superior performance of the Panbio™ RDT was documented in samples with the lowest cycle-threshold RT-PCR values and clients with confirmed clinical symptoms. Thus, we recommend the use of the Panbio™ RDT for both symptomatic and asymptomatic individuals in clinical settings for screening purposes.

## INTRODUCTION

The global public health and economic threats that resulted in countless disastrous effects due to COVID-19 is being revealed in numerous ways and it’s not yet over. More than 6,223,645 deaths from over 504,834,324 documented cases have been reported as of April 18, 2022, since its detection in December 2019 and its being declared as public health emergency in March 2020 [1, 2,3].

In Africa, the current health systems and laboratory diagnostic capacities are at their questionable with respect to managing outbreaks as early as possible. Accordingly, realizing the 2030 SDG with lots of pitfalls in the diagnostic capacity and with so many people failing to get diagnosed. Irrefutably, rapid diagnostic tests are fundamental components of a successful outbreak containment strategy by prompt identification of cases that minimize response costs and save lives due to outbreak of the current COVID-19 [4, 5].

Ethiopia, being one of the countries with limited trained human and material resources is no exception, being the 6^th^ in Africa with 468,895 COVID-19 cases. The country had experienced its fourth wave with an overwhelmingly rapid community transmission compared with the previous epidemic waves, having 35% of positivity rate and 56,706 new cases (Data of December 2021) [3, 6]. Despite this, the country has been tackling the pandemic through allocation of the limited resources for the transmission prevention and implementation of a uniform evidence-based preventive protocol at all levels of the health care system under central command, which is considered as a wise decision for optimal resource utilization [7, 8].

The current choice of established tests for the diagnosis of COVID-19 with a relatively better diagnostic performance using respiratory swab sample is RT-PCR, but its affordability and infrastructure demand for most laboratories in the low-income countries are atypical. Alternative testing modalities have become convenient approaches to reach more clients and a step forward to empower individuals by bringing healthcare services closer to them. In view of this, rapid antigen tests have been in place as an option with their comparable diagnostic performance [9-11].

The impact of rapid diagnostic testing (RDT) has been significant because results can be delivered in short turnaround time of COVID-19 testing. RDT has become a game-changer for triaging patients and crucial medical decisions [7,12]. Compared to RDT, the procedure of RT-PCR is sophisticated, may lead to specimen contamination and technical personnel are prone to acquiring the virus while demand samples from the presumptive cases [4]. On the other hand, several RDTs have been and are currently being developed and commercialized [4, 7,13]. Despite the ease of application and low cost, RDTs are still in need of attention on their quality diagnostic performance for the containment of the virus [14,15].

Since May 2021, Ethiopia has started using Panbio™ COVID-19 rapid antigen test for the diagnosis of COVID-19 in line with RT-PCR [16,17]. Yet, there has been no documented evidence on its diagnostic performance. Indeed, the pandemic urges to have an evidence-based information on its diagnostic accuracy as critically and timely imperative for the program and has its impact in the containment strategy and for continuous quality improvements on its implementation as part of the pandemic control [11,18,19, 20]. Moreover, a lot of studies are available elsewhere that evaluated Panbio™ COVID-19 Ag-RDTs against RT-PCR as the gold standard [21-24], which inevitably leads to bias. Yet, no study has been conducted using BLCM. Thus, this study aimed to assess the diagnostic performance of Panbio™ rapid antigen test for the detection of SARS-CoV-2 in Addis Ababa, Ethiopia using BLCM to bridge the lack of performance accuracy in clinical setting.

## Materials and methods

### Study design, period and settings

A health facility-based prospective cross-sectional diagnostic test evaluation study was conducted from July to September 2021, during on the third wave of the epidemic, among COVID-19 presumptive clients in public health facilities of Addis Ababa, Ethiopia. The study site, Addis Ababa is described in Sisay A. and colleagues,2022 [17]. As part of the current pandemic response, the Addis Ababa Health bureau selected 20 health centers: which is 2 health centers from the ten sub cities, it is based on the previous sub city classification and 6 hospitals for the pilot and start up implementation of this rapid antigen test, Panbio™ (Abbott) rapid antigen test kit for the detection of SARS-CoV-2 using nasopharyngeal swabs against. From these sites, we have selected two government health centers (Kazanches, Kotebe) and two hospitals (Zewuditu Memorial Hospital, Ras Desta Damitew Memorial Hospital). These two public hospitals are amongst the largest and the referral health care system promotes and provides preventive, curative and rehabilitative outpatient care including basic laboratory services [25,26].

### Sampling method

The study population were all presumptive COVID-19 clients among public health facilities of Addis Ababa who were willing to take part in the study and were available during data collection period. We employed a convenience sampling technique until it reached a maximum saturation point (n=438). Stratification of the samples in the four selected sites was done based on the health facilities’ previous three months of SARS-CoV-2 testing performance and availability of resources (see annex 1). The sampling also considered contingency for non-response, invalid and contamination for assuring the best representativeness of the specimens.

### Flow of study Participants

In this study, eligible participants from community surveillance, contacts of confirmed cases, and suspects who fulfill the WHO criteria and Ethiopian guideline for COVID-19 cases were screened by trained professionals as quick triage system [27]. Of these, a total of 438 nasopharyngeal swabs was collected by trained health professionals.

We excluded the critically ill cases, confirmed COVID-19 positive patients and with age ≤ 18 years old. Regarding test kit selection, Panbio™ COVID-19 Ag RDT was used for our study mainly because Panbio™ RDT was the only locally available test kit listed by WHO and authorized by the Ethiopian regulatory body for laboratory utilization [27, 28].

### Methods of Sample collection and data collection instruments

We collected the nasopharyngeal respiratory specimens twice from each study participant upon consent using viral transport medium: one for rapid antigen and the other for RT-PCR. The rapid Ag tests were analyzed immediately according to the manufacturers’ instruction by the primary investigators and results were obtained by the visual interpretation of each testers. The other collected specimen was placed in 3 mL of Viral Transport Medium (VTM) and packed by triple packing system for maintaining the safety measures and shipped immediately to Addis Ababa Public health research and emergency management center laboratory (AAPHEML) for RT-PCR testing. The clinical specimens were collected with strict bio safety measure and lab procedure of Panbio rapid Ag. The quantitative data were collected using a structured data collection tool, prepared was developed in English language after reviewing relevant literature.It included the socio-demographic characteristics and client’s clinical information used for the analysis of the finding of the study [28,29].

### Laboratory Testing Procedures

#### RT-PCR SARS-CoV-2 testing

2 mL VTM (China, Miraclean Technology Co., Ltd., www.mantacc.com) nasopharyngeal specimens were collected and all the nasopharyngeal samples were extracted in BIOER auto method extraction machine with MgaBio plus virus RNA purification kit II and analyzed by using Sansure Biotech (MA-6000) SARS-CoV-2 RT-PCR assay on the BGI Real-Time Fluorescent RT-PCR Kit for Detecting SARS-CoV-2 in a real-time reverse transcription polymerase chain reaction (RT-PCR) test. The assay developed for detecting specific single target gene, which is found on the ORF1ab region of SARS-CoV-2 genome. Further, human housekeeping gene β-Actin is the target gene for the internal control. The master mixing was done by mixing 20μl master mix reagent and 10μl of the extracted sample RNA to the well pre filled with PCR-Mix in the following order: no template (negative) control, patient specimen(s), and positive control. The reference test with a cycle threshold (Ct) < 38 as the criteria for a positive result if Ct value of internal reference not higher than 32 at VIC/HEX and specimen is negative ORF1ab/FAM as 0 or no data available while Ct value at VIC/HEX not higher than 32 [**30]**.

#### Ag-RDT SARS-CoV-2 testing

The collected nasopharyngeal swab was processed on site using the Panbio™ Ag-RDT (Abbott Diagnostic GmbH, Germany). The samples from the swab are mixed with approximately 300 μl of buffer, and then add 5 drops are dispensed into the device. The results were interpreted in 15-20 minutes following in the manufacturer’s instruction but the results do not read after 20 minutes. It detects the presence of the nucleo capsid (N) proteins of the virus on a membrane based using an immune chromatography assay. For a positive result with the Panbio (Abbot) test, a test line must form in the result window (T) and a control (C) line is visible to indicate a test result is valid. In the Negative result is the presence of only the control line(C) and no test line (T) within the result window seen. Invalid result was not occurred in our study if the test line nor the control line was not visible in the result window prior to the specimen dispensed on the device [29]. The test was performed as per Panbio™ COVID-19 Ag Rapid Test Device (NASAL), in vitro diagnostic rapid test for qualitative detection of SARS-CoV-2 antigen (Ag) in a strictly safety percolations [28].

### Data quality Assurance

Data compilers and laboratory workers get its appropriate orientation on how to perform and how to assure valid data using the tool and additional written guide have been provided to them on interpreting each of the study variables. The principal investigators have closely supervised the data collection process so as to ensure the completeness and consistency of the data collection. The laboratory testing was done as per its approved documented standard operating procedures and manufacturer recommendations. In all extraction procedures, as part of assuring the quality management system, we always incorporate a positive and negative quality controls. In addition, data were double entered to prevent error during data entry via cross-checking and also finally checked and verified prior to analysis.

### Data analysis and Interpretation

Descriptive data of the research was entered and analyzed using SPSS statistical software version 23. The rapid antigen test results obtained by visual interpretation were interpreted as positive and negative based on the inbuilt internal control. The RT-PCR laboratory results were interpreted as positive and negative based on the cut-off Ct values of the manufacturer recommendation. All laboratory test results were considered valid if and only if the internal quality control were passed. The binomial 95 % confidence intervals which were obtained following Jeffreys approach in the R package DescTools were used (R version 4.1.3)[and Cohen’s kappa value to assess the agreement beyond chance were obtained with the R package psych [31,34,35]. A value of 1 implies almost perfect agreement and values less than 1 implies less than perfect agreement, with a range of values between 0 and 1 [30,31].

### Bayesian latent class model (BLCM)

With the aim to obtain diagnostic tests accuracies in the absence of a perfect gold standard, Bayesian latent class models (BLCM) were fit to the data following the approach from Hui and Walter for two tests and four populations with MCMC (Markov chain Monte Carlo) simulation to construct posteriors in JAGS version 4.3.0 [32] using the runjags package [33]. The frequencies of the four combinations of dichotomized Panbio and RT-PCR results (++; +-; -+;--) in the four populations, respectively, were modeled with a multinomial distribution. To allow for potential conditional dependencies, pair wise covariance between sensitivities and specificities of all RT-PCRs were included in separate models. Model selection, i.e., in- or exclusion of conditional dependencies was based on the 95% credibility intervals (including 0 or not) and on Bayesian p-values. The model code (S1 File) was obtained with the function “auto huiwalter” of the runjags package [34], with three chains of 50 000 iterations each, a burn-in of 5000 iterations, and a thinning of 10 iterations. Minimally informative priors (beta (1,1)) were used for the sensitivities of both tests, the specificity of Panbio and the four prevalences. The shape parameters for the specificity of the RT-PCR were obtained with beta buster assuming “to be 95% sure that the specificity is greater than 90% with a mode at 99%” as prior information. Convergence was assessed by visual inspection of the trace plots and the potential scale reduction factor (Gelman Rubin statistic) being below 1.1. A sensitivity analysis was performed by using different combinations of minimally (dbeta(1,1)) or weakly informative priors (dbeta(2,1))

### Ethical consideration

Ethical approval was obtained from IRB of department of medical laboratory Sciences, College of Health Sciences, Addis Ababa University (reference-MLS/174/21)), IRB office of Health bureau, AAPHREML (Reference-AAHB/4039/227) and also from Addis Ababa University, College of natural and computational science IRB (IRB-CNCSDO/604/13/2021). Additionally, AAPHREML wrote support letter to the study health facilities. During data collection process the data collectors informed each study health facility and study participants about the purpose and anticipate benefits of the research and on their full right to refuse, withdraw or completely reject part or all of their part in the study. Written informed consent on the use of data with full anonymity was obtained from the voluntary participants. This work has been done and performed as per Helsinki declaration.

## Results

### Socio-demographic characteristics of the study variables of participants

A total of 438 presumptive clients were identified and enrolled in this study and the majority of them, 239 (54.6%) were females and the age of the participants were ranging from 18 to 84 years and the mean age was 36.38 ±14.3 years. Three fourth of the study participants (n=318) had symptoms of COVID-19 and the most reported clinical symptoms were cough (n=191), followed by headache (n=39). For more than half (n=258) of the participants, the reason for getting tested was due to observing the classic symptoms. The detail demographic data were depicted in table 1.

**Table 1.**
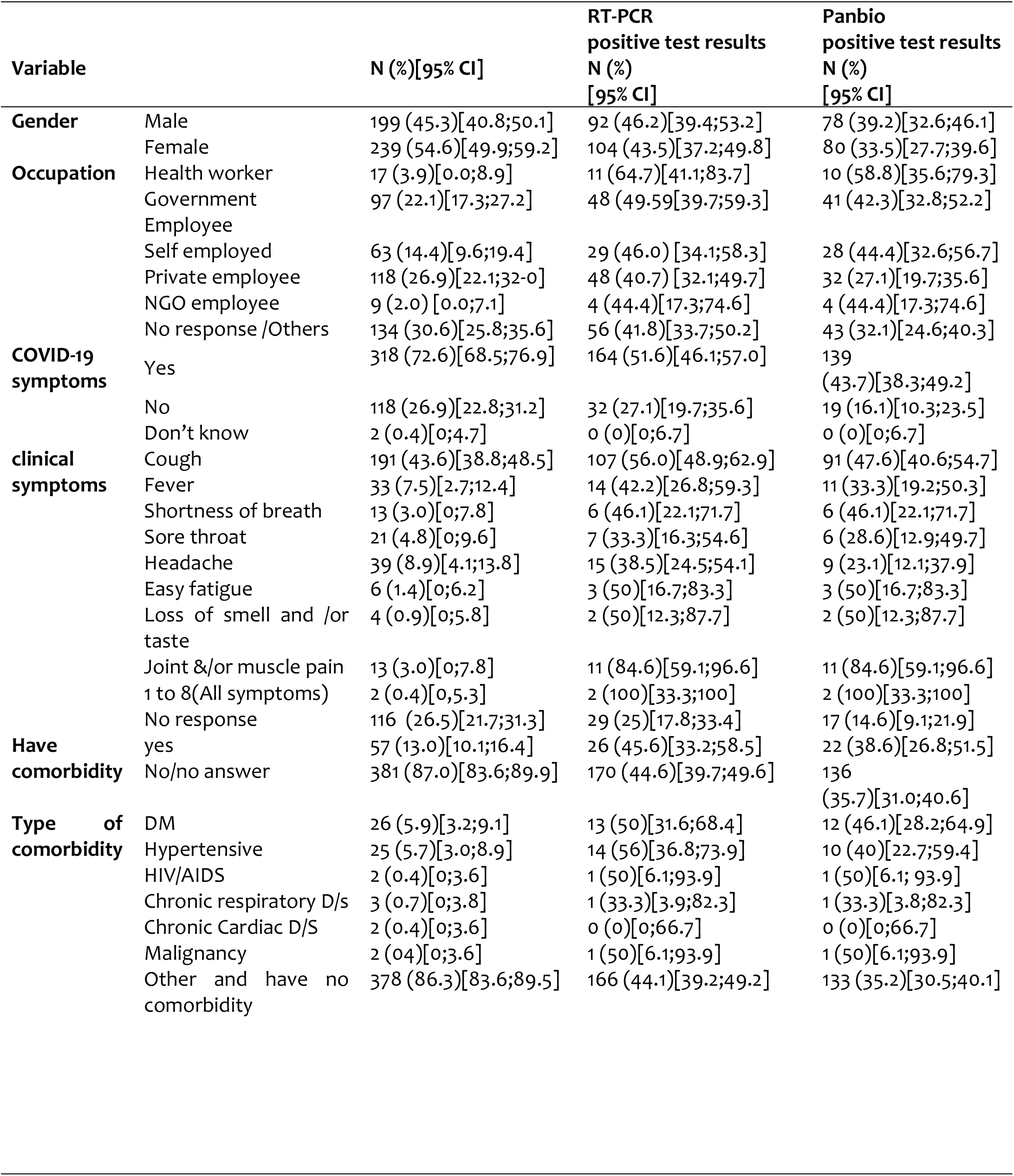
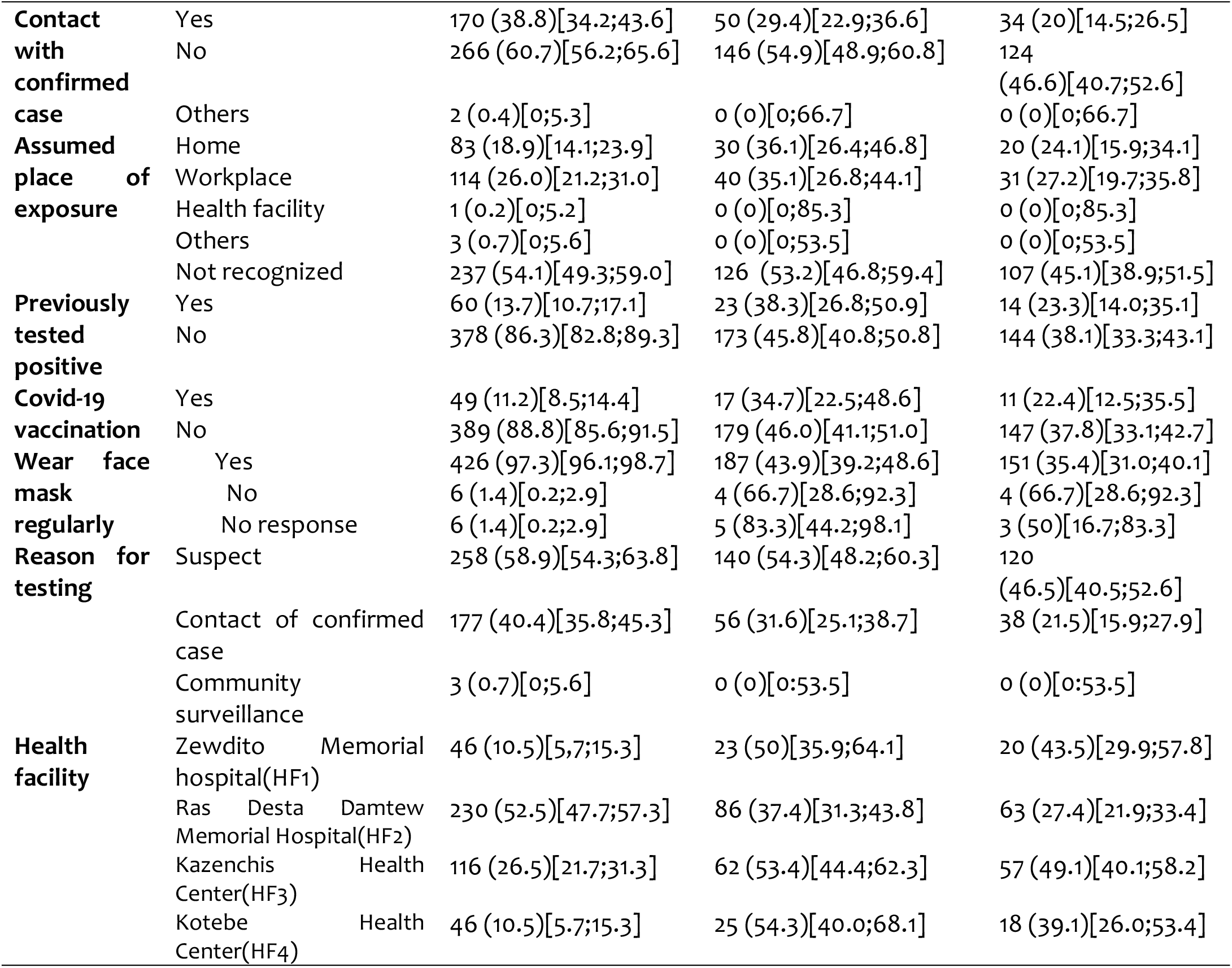
Demographic characteristics of study participants and cross-classified results of RT-PCR and Panbio, Addis Ababa, Ethiopia, 2021 (n=438)

### Test performance accuracy based on days since Clinical symptom onset

The performance accuracy of the RDT were high as of the lowest number of ct values as depicted in table 2. As the ct values of the test Sensitivity Performance of the Rapid Antigen Test Kits with the Date of Clinical Onset of Symptoms of the Clients, and ct values 2022, Addis Ababa, Ethiopia

**Table 2.**
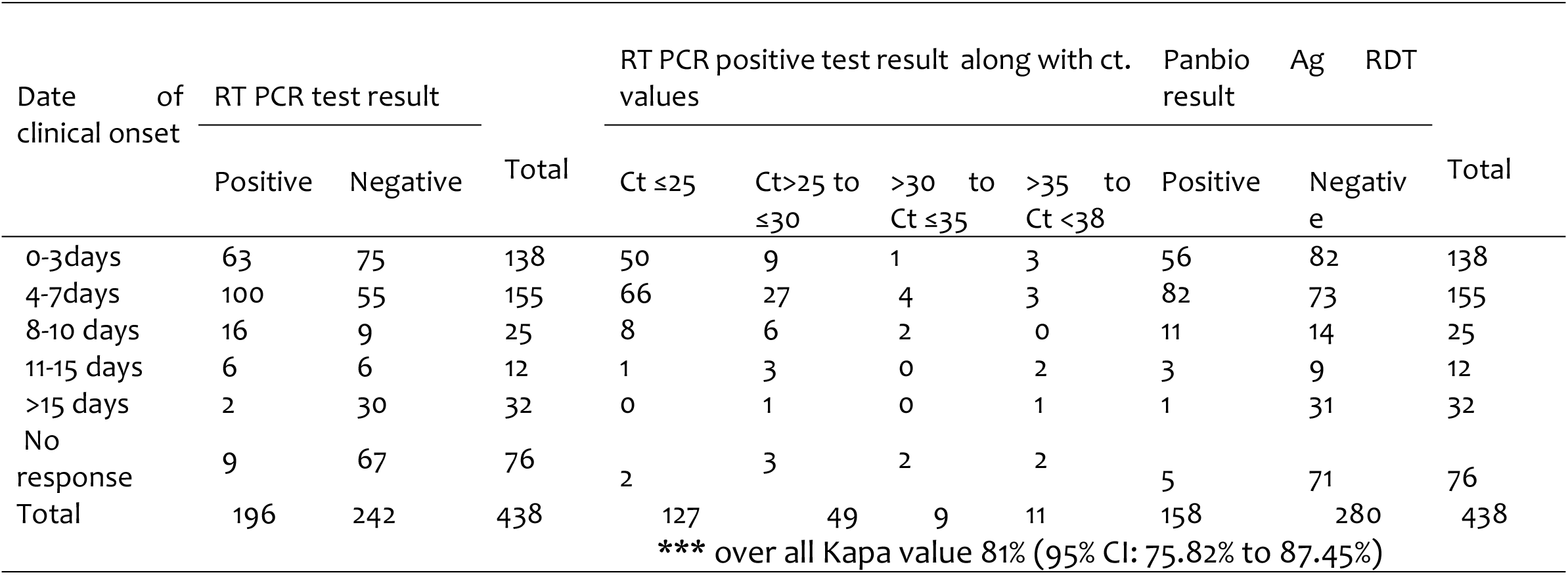
Test performance along date of clinical onset and its CT values, 2021, Addis Ababa, Ethiopia.

### Diagnostic Performance of Panibo Ag test and RT-PCR using BLCM

We have performed BLCMs to assess the diagnostic performance of the Panbio™ antigen test and RT-PCR in the absence of a gold standard using 4 population with 2 tests, with symptoms and without symptoms using BLCM. Accordingly, the Ag-RDT and RT-PCR had a sensitivity (95% CrI) of 99.6 %(98.4%-100 %), 89.3 %(83.2%-97.6%) and specificity (95% CrI) of 93.4% (82.3% - 100 %), 99.1 %(97.5%-100%) respectively, table 3a.

**Table 3a.**
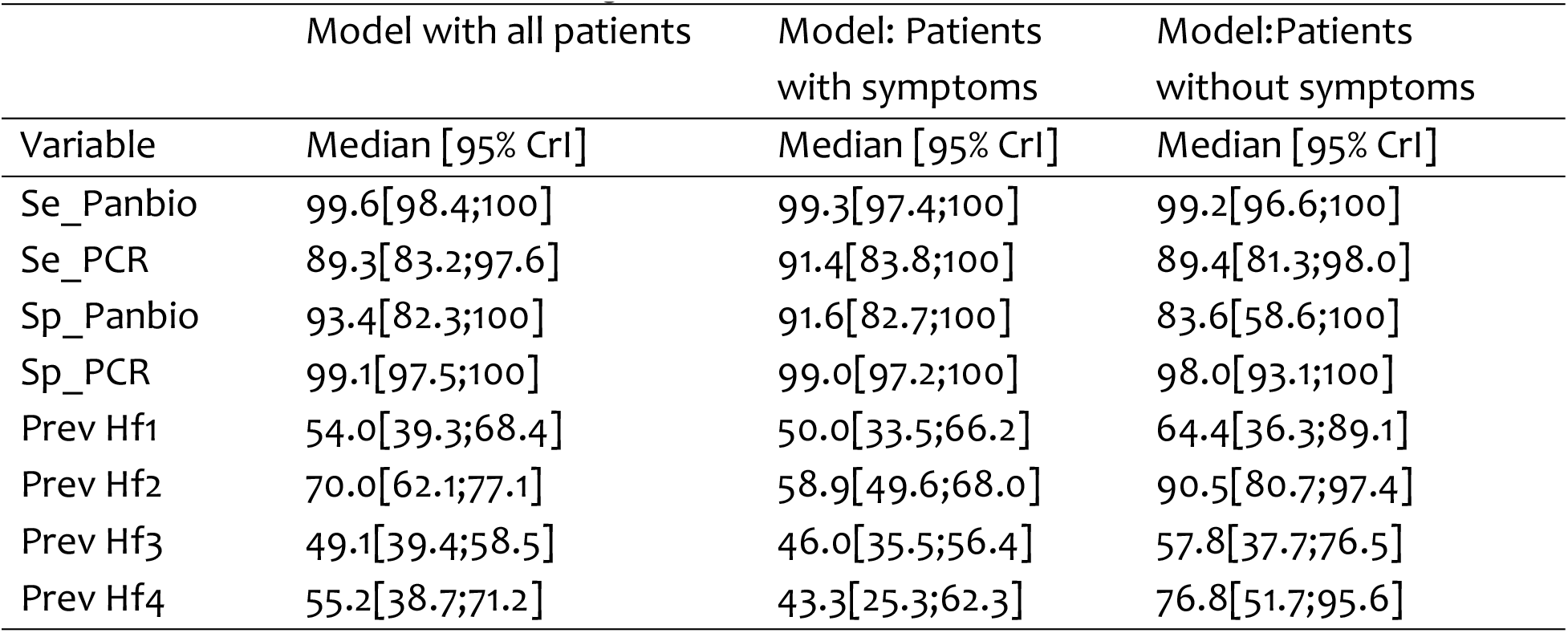
Performance of test kits using all model of BLCM, Addis Ababa, Ethiopia, 2021

The performance of Ag-RDT was also examined using different parameters or conditional dependency, as depicted in table 3b and S1-S6 (supplementary files, S1-S6).

**Table 3b:**
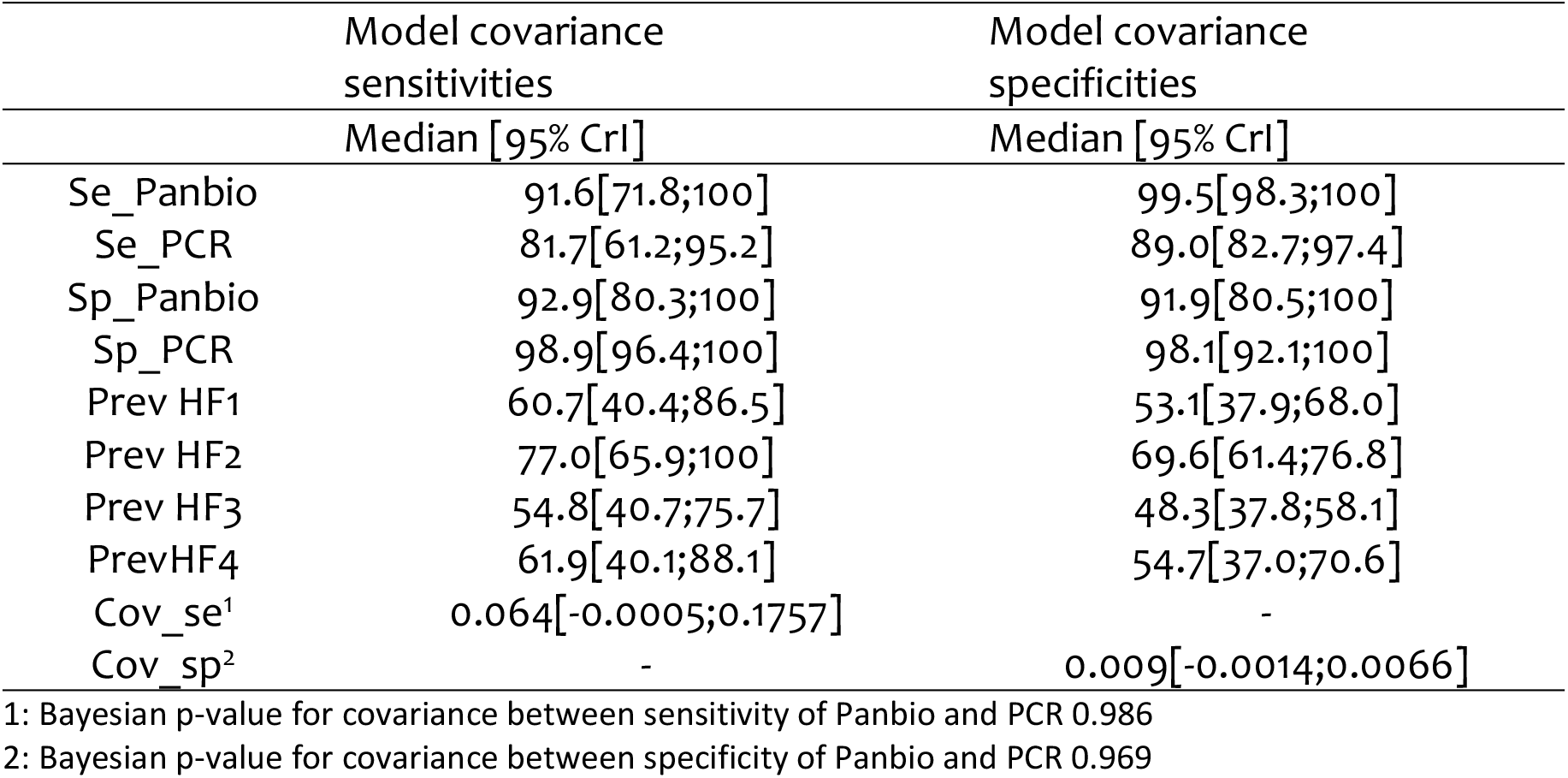
Performance of test kits on conditional dependency using BLCM, Addis Ababa, Ethiopia, 2021

## Discussion

Accurate reliable rapid diagnostic tests play a very crucial role in curbing COVID-19 infections. Accordingly, rapid Ag-based diagnostic tests that could pinpoint patients at early disease were commonly recommended. In this regard, the current status of our country’s laboratory diagnosis system for COVID-19 is centralized with limited number of facilities resulted in longer turnaround of results, which goes against the escalation of the virus in a country having more than 100 million population and not more than 4,000 diagnostic medical laboratories [7, 16].

The diagnostic sensitivity and specificity of Panbio was comparable with reports by Nsoga *et. al*. with sensitivity and specificity ranges of 71.4%-91.7% and 94.9%-100%, respectively, while using RT-PCR as a gold standard. These study results also considered Ct values of <30 that yielded test sensitivities from 87.7% to 97.8% [36-38].

In terms of predictive values: when we use RT-PCR as gold standard, our finding had comparable PPV and incomparable NPV when compared and contrasted with a study by Bulilete et al., with the overall sensitivity of 71.4% (95% CI: 63.1%, 78.7%), the specificity of 99.8% (95% CI: 99.4%, 99.9%), the positive predictive value of 98.0% (95% CI: 93.0%, 99.7%) and a negative predictive value of 96.8% (95% CI: 95.7%, 97.7%) [39].This indicates a clear difference in the specificity of the test performance among the negative cases, which may be due to the difference in study participant recruitments, which is we used more the asymptomatic and having clients with chief compliant of COVID-19 while presenting in the health facilities [39].

Our study finding illustrate more inline finding with the WHO stated criteria for the emergency use of clinical setting of such products as appropriate criteria, such as WHO’s priority target product profiles for COVID-19 diagnostics of acceptable’ sensitivity ≥ 80% and specificity ≥ 97%), can be considered as a replacement for laboratory-based RT-PCR when immediate decisions about patient care. This is truer in symptomatic clients and we are not sure that this performance be in asymptomatic cases. Indeed, Panbio™ COVID-19 Ag-RDTs has a similar sensitivity performance in both symptomatic and asymptomatic, but its specificity is lower among clients without symptoms. Whereas RT-PCR is slightly lower in clients among without symptoms [40, 41].

As a general principle RDTs can be used outside laboratories, at/ or near the point of care. They are easy to use, provide rapid results and do not require any expensive equipment. Ag-RDTs can be considered as alternatives to expand NAAT here (NAAT) for direct detection of SARS-CoV-2 virus for diagnosis of early COVID-19 and also its critical tool to scale-up testing and diagnostic in the fight against COVID-19 for the control of the pandemic. Nevertheless, we highly underline that these tests should be performed under strict follow-up of trained professional starting from the very beginning of pre analytical to post analytical of result dissemination [42].

A similar study conducted in Germany, Switzerland and Japan in comparing this RDT diagnostic performance and using RT-PCR as a gold standard method of the classical method model showed more comparable finding among symptomatic study participants and its superior performance have been observed in case of samples having the lowest ct values and high amount of viral concentration. We understand that, there might be a direct relationship with the ct values and viral load of the study samples. However, because of resource limitation, we did not monitor the viral concentration of the clients and readers should consider it while inferring our finding with these studies [12, 15, 43, 44].

Our finding revealed the Panbio™ rapid antigen test have highest level of agreement of the inter-test agreement beyond chance of the established assays (RT-PCR) with an average Cohen’s kappa value of 81% (95% CI: 75.82% to 87.45%) among our study subjects of 438 COVID-19 suspected individuals, which was consistent with similar studies [36, 37] and lower finding compared with Torres et. al., where the defined study population was different form the present study [45].

The diagnostic performance Panbio™ Covid-19 Ag rapid test highly correlate with Ct values and day of clinical onset. We found a superior performance in lowest ct values, which is most probably during at this condition the viral concentration become high and as the clinical onset of the day increase its performance among asymptomatic cases [37, 41].

Most of the false negative Ag-RDT results were among the population with contacts of SARS-CoV-2 confirmed patient within a week range of contact of confirmed case and they have reported no clinical symptoms and chief complaints when arriving at the testing health facilities. Their age ranged from 18 to 68 years old and most of them were female. Also there was a single false positive report from the total non-diseased of 242. The low frequency of false positive report of the AG-RDTwas in line with a study done by Bulilete and colleagues in Spain and Akingba and colleagues, in south Africa, while we consider RT-PCR as gold standard [38,39].

This study proved the presence of a clear difference in the performance of the test kits when taking RT-PCR as a perfect gold standard, instead of using BLCM assuming no gold standard.

## Conclusion and recommendations

The introduction of rapid antigen laboratory diagnostic methods in routine laboratory for identification of SARS-COV-2 for a possibility of expanding to the general service, especially near to the patients which could help medical practitioners for isolating the patients and intervention of infections that can help to combat the devastating transmission. Based on our study findings, the diagnostic performance of the Panbio™ Covid-19 RDT were 99.6 % sensitivity and 93.4 % specificity among presumptive cases of Addis Ababa, by which it qualify the WHO established criteria required to have for Ag-RDTs (sensitivity ≥80% and specificity ≥97%). Thus, it’s highly recommended to use it in areas of symptomatic individuals. This can minimize reoccurrence and further spread of the pandemic and gripped the spread of the virus by testing all the presumptive. We highly recommended further large-scale study coupled with genomic approaches considering the presence of performance variation as the variant of concern continues to emerge.

## Data Availability

All data set used in this study are included in this manuscript.

## Supporting information

S1 –S6 are supplementary material used for this BLCM analysis, Addis Ababa, Ethiopia, 2022.

## Authors’ contributions

**Conceptualization:** Abay Sisay, Adey Feleke Desta

**Data curation**: Abay Sisay, Abebaw Tiruneh, Yasin Desalegn, Abraham Tesfaye, Adey Feleke Desta

**Formal analysis:** Abay Sisay, Sonja Hartnack,

**Investigation:** Abay Sisay, Sonja Hartnack, Abebaw Tiruneh, Yasin Desalegn, Abraham Tesfaye, Adey Feleke Desta

**Methodology:** Abay Sisay, Sonja Hartnack, Abebaw Tiruneh, Abraham Tesfaye, Adey Feleke Desta

**Project administration**: Abay Sisay,

**Resources:** Abay Sisay,

**Software:** Abay Sisay, Sonja Hartnack, Adey Feleke Desta

**Supervision:** Abay Sisay, Sonja Hartnack, Abebaw Tiruneh, Abraham Tesfaye, Adey Feleke Desta

**Validation:** Abay Sisay, Sonja Hartnack, Abebaw Tiruneh, Abraham Tesfaye, Adey Feleke Desta

**Visualization:** Abay Sisay, Sonja Hartnack, Adey Feleke Desta

**Writing original draft:** Abay Sisay, Sonja Hartnack

**Writing review & editing**: Abay Sisay, Sonja Hartnack, Abebaw Tiruneh, Yasin Desalegn, Abraham Tesfaye, Adey Feleke Desta

## Data Availability Statement

All data set used in this study are included in this manuscript.

## Acknowledgments

We are very grateful and acknowledge the Addis Ababa University and Addis Ababa Health bureau, the Zewditu Memorial Hospital, the Ras Desta Damtew Memorial Hospital, the Kotebe Health Center and the Kazanches Health Center for granting their institutional ethical approval and their strong support and assistance in accessing diverse resources used in the study.

We also acknowledge all the data collector and study participants.

## Supplementary materials, S1-S6 (supplementary files, S1-S6)

S1: BLCM. Code. R

S2: performance analysis with weakly informative priors

S3: Graphical output from the final BLCM (trace plot, ECDF, density and autocorrelation plot) for sensitivity of Panbio rapid antigen test

S4: Graphical output from the final BLCM (trace plot, ECDF, density and autocorrelation plot) for sensitivity of RT-PCR

S5: Graphical output from the final BLCM (trace plot, ECDF, density and autocorrelation plot) for specificity of Panbio rapid antigen test

S6: Graphical output from the final BLCM (trace plot, ECDF, density and autocorrelation plot) for specificity of RT-PCR

**Annex 1.** Stratification of the samples in the four selected sites we allocate the total proposed 438 sample in to the four selected sites accordingly.

| Health Facility name | Assigned code | Assigned sample size |
| --- | --- | --- |
| Zewditu memorial Hospital | 01 | 46 |
| Ras Desta Damtew memorial Hospital | 02 | 230 |
| Kazanchis Health Ceneter | 03 | 116 |
| Kotebe Health Center | 04 | 46 |

